# The variation of malaria prevention measures knowledge and their associated factors: A cross-sectional study in East Nusa Tenggara Province Indonesia

**DOI:** 10.1101/2023.09.12.23295402

**Authors:** Robertus Dole Guntur, Maria A. Kleden, Damai Kusumaningrum, Fakir M Amirul Islam

## Abstract

**Objective:** Malaria is one of the major public health problems in African and Southeast Asian countries including Indonesia. However, knowledge of malaria prevention measures (MPM) is not well studied, particularly in Indonesia. This study aimed to investigate the level of MPM knowledge and associated factors among rural adults in high, moderate and low endemic settings of East Nusa Tenggara Province (ENTP) Indonesia.

**Methods:** A community-based cross-sectional study was conducted among a randomly selected 1495 households at rural ENTP. Multistage sampling technique was employed to recruit participants. Univariate and multivariable logistic regression model was used to assess factors affecting knowledge of MPM.

**Results:** The level of MPM knowledge in low, moderate, and high endemic settings differs significantly with the highest in low settings (57%, 95% CI: 50.5 – 63.5 and the lowest in high settings (19.3%, 95% CI: 11.1 – 27.5). In all settings, good level of MPM knowledge was significantly higher for adults with high SES (Adjusted odds ratio (AOR) = 2.52, 95% CI: 1.20 – 5.30; AOR=20.5,95%CI: 4.64-90.8, AOR=3.31,95%CI: 1.34-8.15 respectively) compared to those having low SES. In high and moderate settings, the likelihood of good MPM knowledge was considerately higher for adults with at least secondary (AOR=2.35, 95% CI= 1.29 – 4.36, AOR=2.66, 95% CI=1.32-5.39 respectively) than those with primary or no education level.

**Conclusions:** The good level of MPM knowledge was very low in three different malaria endemic settings. Higher level of education and high SES were significantly associated with the good level. Therefore, health education promotion on MPM knowledge is critical to support malaria elimination program in the province.

## Introduction

Malaria is a communicable disease spreading across 84 countries globally (1). The World Health Organization (WHO) projected that the number of malaria cases was about 247 million in 2021 worldwide with 95% of the cases was contributed by African region (1). In South East Asia, the number of malaria cases was predicted about 5,383,185 in 2021, of which 79% were from India and 15% were from Indonesia (1). The number of malaria cases in Indonesia shows an increasing trend with the highest burden of malaria was in the Eastern part of the country(2). Most of the districts in the Western part of Indonesia have been classified as malaria elimination area, whilst it was limited in the Eastern part of the country (2).

ENTP which is one of the lag province in the Eastern part of Indonesia (3) was the third-highest contributor of the malaria burden to the nation in 2021 (2). The province has 22 districts of which 13.6 % of the total number of district were classified as high MES, 63.6% was low MES, and 22.7% was malaria free zone (2). Under the partnership with the National Malaria Control Program of Indonesia government, local authority of ENTP has applied various effort to reduce the burden of malaria in this region. This includes ensuring the availability of artemisinin-based combination therapies (ACTs) for malaria treatment in all local health facilities(2), increasing coverage of ACT from 55% in 2013 (4) to 99.8% in 2021(2), conducting indoor residual spraying (IRS) to respond outbreak, larva control and environmental management (5,6), increasing coverage of mass distribution of insecticide treated mosquito nets from 9 districts in 2010 (6) to 15 districts in 2017 (7). However, the number of malaria cases are still high with the total number of cases in 2021 was 9,419 cases (2).This implies that these approaches might not enough and the implementation of those measures might depend on the community behaviour which is less documented in the study area. To progress to malaria elimination, active participation of community is critical (8) and to do that the community should have malaria awareness including knowledge in malaria prevention measures (MPM). Having high level of MPM knowledge leads to high practice of MPM (9) and high participation in various malaria elimination programs which in turn to speed up malaria elimination (10).

Many studies on malaria prevention measures at the global level indicated that the knowledge of malaria prevention measures was significantly associated with the education level of participants (11–13). A recent systematic review on MPM in the society of Southeast Asia nations implies that the studies of MPM and its associated factors was restricted in the zone (14). To boost malaria elimination, local knowledge of malaria prevention measures should be measurable and the level of malaria prevention knowledge of the local population should be integrated in designing malaria elimination programmes (14).

In Indonesia, limited research on the community-level MPM has been performed (15–17). A study of malaria prevention knowledge of rural adults in Purworejo district of Central Java showed that about half of the participants had knowledge of mosquito nets and covering ventilation for preventing malaria, and the usage of these measures was considerably associated with the education of participants (15). A study on 50 villages in Central Java shows that more than half of 1000 rural communities kept their house clean and applied indoor residual spray to prevent malaria (16). However, all of these investigations were carried out in the Western part of Indonesia, which most of the districts in this area have been classified as malaria free zone. The recent study in the Eastern part of Indonesia, including in ENTP, indicated that the practice of various types of MPM differs amongst provinces (17).

Some studies on MPM had been conducted in the context of rural ENTP (18–22) A study on knowledge of LLINs and non-LLINs for rural community for this province indicated that rural adults in low endemic settings was knowledgeable with non-LLINs, whilst it was LLINs in high endemic settings (18). Another study on Tetun ethnicity in Timor island indicated that most of community in that group had knowledge on the traditional plants for preventing malaria (19). Other studies recruiting 1503 participants rural adults from 49 villages in ENTP reveals that the level of knowledge in some kinds of MPM of rural population was poor (20–22), however the disparity of MPM knowledge amongst different malaria endemic settings has not been investigated in those studies. Moreover, the discrepancy of level of MPM knowledge amongst different settings and its associated factors have not been explored this study area. The local community should be able to identify various kinds of MPM and their knowledge of MPM should be considered in planning and implementing malaria elimination programs to succeed the program (14). The high level of knowledge of MPM leads to a high level of practice of MPM in their daily life (9), and the combination of some kinds of MPM tailored to the local population would boost malaria elimination (23–25). Furthermore, understanding the knowledge of MPM in the local community and identifying which groups are the most vulnerable in the population will help local authorities design sustainable malaria programs to accelerate malaria elimination. Therefore, this study investigated the discrepancy in MPM knowledge amongst different malaria endemic settings and its associated factors. It is anticipated these findings would enhance the expectation of ENTP and the Indonesian government to achieve a malaria elimination zone by 2030 (26).

## Materials and Methods

### Study settings

This community based cross-sectional study was performed in 3 out of 22 districts in the province from October to December 2019. Firstly, East Sumba district which is 52.3% population of the district working in agricultural sector was classified as high malaria endemic setting (27,28). The area of the district is 7,000.50 km^2^, with a population density of 35 people per square kilometer (29). Secondly, Belu district, which is 37.7% population of the district working in agricultural sector was classified as moderate malaria endemic setting (27,28). The area of the district is 1,284.94 km^2^, with a population density of 177 people per square kilometer (30). Finally, East Manggarai district with 78.8% population of the district working in agricultural sector was classified as low malaria endemic setting (27,28). The area of the district is 2,401.39 km^2^, with a population density of 111 people per square kilometer (31)

### Study population and sample size calculation

All adults in three selected malaria endemic settings were study population for this research. In total, the number of respondents enrolled for this study was 1503 adults. This number was obtained after considering of malaria prevalence in ENTP, intra-class correlation of malaria prevention study in Indonesia, design effect and participation rate of participants. The comprehensive computation of sample size was reported on the previous article of the authors (32).

### Data collection procedure

Data collection tools was adapted from validated questionnaire (33) with some modifications. Originally the questionnaire was prepared in English. Then, the local language expert and the main author of this study translated the questionnaire into local language (Indonesian). They then combined the translation version of the questionnaire. The combined version of the questionnaire was used as a final tool to collect data. Nine enumerators having background in nursing school and had working experience in the local public health centres were employed and educated about the aim of the study including overview of malaria prevention measures knowledge, how to approach participants and obtain their written consent, how to complete the questionnaire. Data collection was supervised strictly by the main author of this study. The completeness of the questionnaire was monitored on daily basis and the incomplete questionnaire was returned to enumerators on the following day for correction by re-visiting the household.

### Study variables and operational definitions

The dependent variable of the study was the level of malaria prevention measures (MPM) knowledge in each malaria endemic settings. The good level of MPM knowledge was defined based on the response of participant on six questions. It is including questions related to sleeping under LLINs, sleeping under non-LLINs, keeping surrounding house clean, using mosquito coil, wearing long sleeved clothes when go outdoors at night, using Indoor Residual Spraying. Each of question has option yes or no, with yes obtaining of score one. Therefore, the total score of each participant ranged from zero to six. Participants who could answer correctly at least three question related knowledge of malaria prevention measures were categorized as having a good knowledge of MPM, whilst participants answering zero to two questions were categorized as having a poor knowledge of MPM (12).

The independent variables of the study were gender, age group, education level, occupation, family size, socio-economic status (SES), household income, the nearest health facilities, the distance to the nearest health facilities, and the location of household. In this study gender was categorized as male and female, age group was classified as less than 30 years old, 30-39 years, 40-49 years, 50-59 years, 60 years or above. The education level was classified into two categories: primary school or less and secondary school or above, the main occupation was classified into four groups: farmer, housewife, office staff and others. The location of household was categorized as coastal, hills and other areas. The SES group of participants was classified as low, average and high.

### Data Analyses

Socio-demographic data of participants was described by descriptive statistics. The proportion of participants answering correctly for each question of malaria prevention measures and its 95% confidence interval (CI) was computed in each malaria endemic settings. The percentage of good knowledge for each malaria endemic settings was calculated with its 95% CI. To investigate the potential factors affecting good knowledge of malaria prevention measures, a univariate and multivariate binary logistic regression analysis was applied. Adjusted odds ratio with 95% CI and p value less than 5% was employed to confirm the significance of each variable.

### Ethics approval and consent to participate

This study was accepted by Human Ethics Committee of the Swinburne University of Technology (reference number 20191428-1490) and the Indonesian Ministry of Health (reference letter: 164 LB.02.01/2/KE.418/2019). Permission letter was further obtained from the governor of ENTP, head of East Sumba, Belu, and East Manggarai district, nine head of sub-districts, and forty-nine village leaders in this region. Information related to the purposes, risk, and advantage of the study was provided to all participants prior to data collection. Participation in this study was fully voluntary and written consent was attained from each participant.

## Results

### Distribution of participants having awareness that malaria could be prevented by socio-demography characteristics

The distribution of participants having awareness that malaria could be prevented was shown in Table 1. Most of participants in high malaria endemic settings (94.1%) had awareness that malaria could be prevented, whereas it was only 77.6% and 72.4% in low and moderate endemic setting respectively. In all malaria endemic settings, the awareness that malaria could be prevented was in line with the increase education level of participants. In terms of socio-economic status (SES) of participants, the highest proportion of participants having awareness malaria could be prevented was from the high SES group both for rural adults in moderate and high malaria endemic settings with 98.4% and 96.6% respectively, whilst it was from low SES group in low endemic settings.

**Table 1:** Distribution of participants having awareness that malaria could be prevented by socio-demographic factors in each malaria endemic settings (N=1495.

| Characteristics | ENTP |  | Malaria Endemic Settings |  |  |  |  |  |
| --- | --- | --- | --- | --- | --- | --- | --- | --- |
|  | Yes, n (%) | Total | High |  | Moderate |  | Low |  |
|  |  |  | Yes, n (%) | Total | Yes, n (%) | Total | Yes, n (%) | Total |
| **Overall** | 1216<br>(81.3) | 1495 | 466<br>(94.1) | 495 | 362<br>(72.4) | 500 | 388<br>(77.6) | 500 |
| **Gender** |  |  |  |  |  |  |  |  |
| Male | 614<br>(84.5) | 727 | 216<br>(93.5) | 231 | 190<br>(81.5) | 233 | 208<br>(79.1) | 263 |
| Female | 602<br>(78.4) | 768 | 250<br>(94.7) | 264 | 172<br>(64.4) | 267 | 180<br>(75.9) | 237 |
| **P-Value** | 0.003 |  | 0.574 |  | < 0.001 |  | 0.401 |  |
| **Age group** |  |  |  |  |  |  |  |  |
| < 30 | 177<br>(86.3) | 205 | 75<br>(94.9) | 79 | 54 (84.4) | 64 | 48<br>(77.4) | 62 |
| 30 -39 | 362<br>(86.6) | 418 | 134<br>(97.8) | 137 | 93 (86.1) | 108 | 135<br>(78.0) | 173 |
| 40 - 49 | 313<br>(84.4) | 371 | 133<br>(96.4) | 138 | 93 (75.6) | 123 | 87<br>(79.1) | 110 |
| 50 - 59 | 221<br>(74.9) | 295 | 66<br>(95.7) | 69 | 79 (61.2) | 129 | 76<br>(78.4) | 97 |
| > 60 | 143<br>(69.4) | 206 | 58<br>(80.6) | 72 | 43 (56.6) | 76 | 42<br>(72.4) | 58 |
| **P-Value** | < 0.001 |  | < 0.001 |  | < 0.001 |  | 0.896 |  |
| **Level of education** |  |  |  |  |  |  |  |  |
| Primary school or less | 731<br>(76.2) | 959 | 351<br>(92.9) | 378 | 182<br>(61.1) | 298 | 198<br>(70.0) | 283 |
| Secondary School or Above | 485<br>(90.5) | 536 | 115<br>(98.3) | 117 | 180<br>(89.1) | 202 | 190<br>(87.6) | 217 |
| **P-Value** | < 0.001 |  | 0.029 |  | < 0.001 |  | < 0.001 |  |
| **Main Occupation** |  |  |  |  |  |  |  |  |
| Farmer | 690<br>(83.0) | 831 | 321<br>(94.4) | 340 | 115<br>(70.1) | 164 | 254<br>(77.7) | 327 |
| Housewife | 284<br>(70.5) | 403 | 80<br>(90.9) | 88 | 151<br>(63.7) | 237 | 53<br>(67.9) | 78 |
| Other | 102<br>(92.7) | 110 | 48<br>(98.0) | 49 | 43 (97.7) | 44 | 11<br>(64.7) | 17 |
| Office staf | 140<br>(92.7) | 151 | 17<br>(94.4) | 18 | 53 (96.4) | 55 | 70<br>(89.7) | 78 |
| **P-Value** | < 0.001 |  | 0.390 |  | < 0.001 |  | 0.006 |  |
| **Socio-Economic Status** |  |  |  |  |  |  |  |  |
| Low | 336<br>(74.8) | 449 | 139<br>(92.1) | 151 | 41 (39.0) | 105 | 156<br>(80.8) | 193 |
| Average | 710<br>(82.6) | 860 | 271<br>(94.8) | 286 | 258<br>(77.9) | 331 | 181<br>(74.5) | 243 |
| High | 170<br>(91.4) | 186 | 56<br>(96.6) | 58 | 63 (98.4) | 64 | 51<br>(79.7) | 64 |
| **P-Value** | < 0.001 |  | 0.368 |  | < 0.001 |  | 0.263 |  |
| **Family size** |  |  |  |  |  |  |  |  |
| < = 4 | 633<br>(78.8) | 803 | 190<br>(91.3) | 208 | 260<br>(72.4) | 359 | 183<br>(77.5) | 236 |
| > 4 | 583<br>(84.2) | 692 | 276<br>(96.2) | 287 | 102<br>(72.3) | 141 | 205<br>(77.7) | 264 |
| **P-Value** | 0.007 |  | 0.024 |  | 0.985 |  | 0.977 |  |
| **The nearest Health Service** |  |  |  |  |  |  |  |  |
| Village maternity posts | 310<br>(80.3) | 386 | 131<br>(89.1) | 147 | 166<br>(74.8) | 222 | 13<br>(76.5) | 17 |
| Village health Post | 217<br>(71.9) | 302 | 34<br>(100.0) | 34 | 56 (53.8) | 104 | 127<br>(77.4) | 164 |
| Subsidiary Public Health centres | 290<br>(85.8) | 338 | 168<br>(97.1) | 173 | 35 (64.8) | 54 | 87<br>(78.4) | 111 |
| Public Health centres | 399<br>(85.1) | 469 | 133<br>(94.3) | 141 | 105<br>(87.5) | 120 | 161<br>(77.4) | 208 |
| **P-Value** | < 0.001 |  | 0.009 |  | < 0.001 |  | 0.996 |  |
| **Distance to the nearest health service** |  |  |  |  |  |  |  |  |
| < 1 Km | 460<br>(79.6) | 578 | 165<br>(94.8) | 174 | 177<br>(65.3) | 271 | 118<br>(88.7) | 133 |
| 1 - 2 Km | 328<br>(82.0) | 400 | 84<br>(96.6) | 87 | 121<br>(81.2) | 149 | 123<br>(75.0) | 164 |
| >=3 Km | 428<br>(82.8) | 517 | 217<br>(92.7) | 234 | 64 (80.0) | 80 | 147<br>(72.4) | 203 |
| **P-Value** | 0.368 |  | 0.386 |  | 0.001 |  | 0.001 |  |
| **‡HH Income in relation to PMW** |  |  |  |  |  |  |  |  |
| < PMW | 1082<br>(80.6) | 1342 | 431<br>(93.9) | 459 | 327<br>(71.7) | 456 | 324<br>(75.9) | 427 |
| >= PMW | 134<br>(87.6) | 153 | 35<br>(97.2) | 36 | 35 (79.5) | 44 | 64<br>(87.7) | 73 |
| **P-Value** | 0.036 |  | 0.414 |  | 0.267 |  | 0.026 |  |
| **Location of household** |  |  |  |  |  |  |  |  |
| Coastal Area | 186<br>(91.6) | 203 | 85<br>(97.7) | 87 | 78 (97.5) | 80 | 23<br>(63.9) | 36 |
| Others | 251<br>(82.0) | 306 | 123<br>(94.6) | 130 | 43 (62.3) | 69 | 85<br>(79.4) | 107 |
| Hills | 779<br>(79.0) | 986 | 258<br>(92.8) | 278 | 241<br>(68.7) | 351 | 280<br>(78.4) | 357 |
| **P-Value** | < 0.001 |  | 0.229 |  | < 0.001 |  | 0.120 |  |

### Knowledge of malaria prevention measures of participants

The variation of knowledge of malaria prevention methods of rural adults in different malaria endemic settings was presented in Table 2. Overall, there was a discrepancy of good level of malaria prevention measures knowledge in these settings with the highest level was in low malaria endemic settings at 57.0% with 95% confidence interval (CI): 50.5 – 63.5. Whilst the lowest level of good knowledge of MPM was in high malaria endemic settings at 19.3% with 95% CI: 11.1 – 27.5.

**Table 2.**
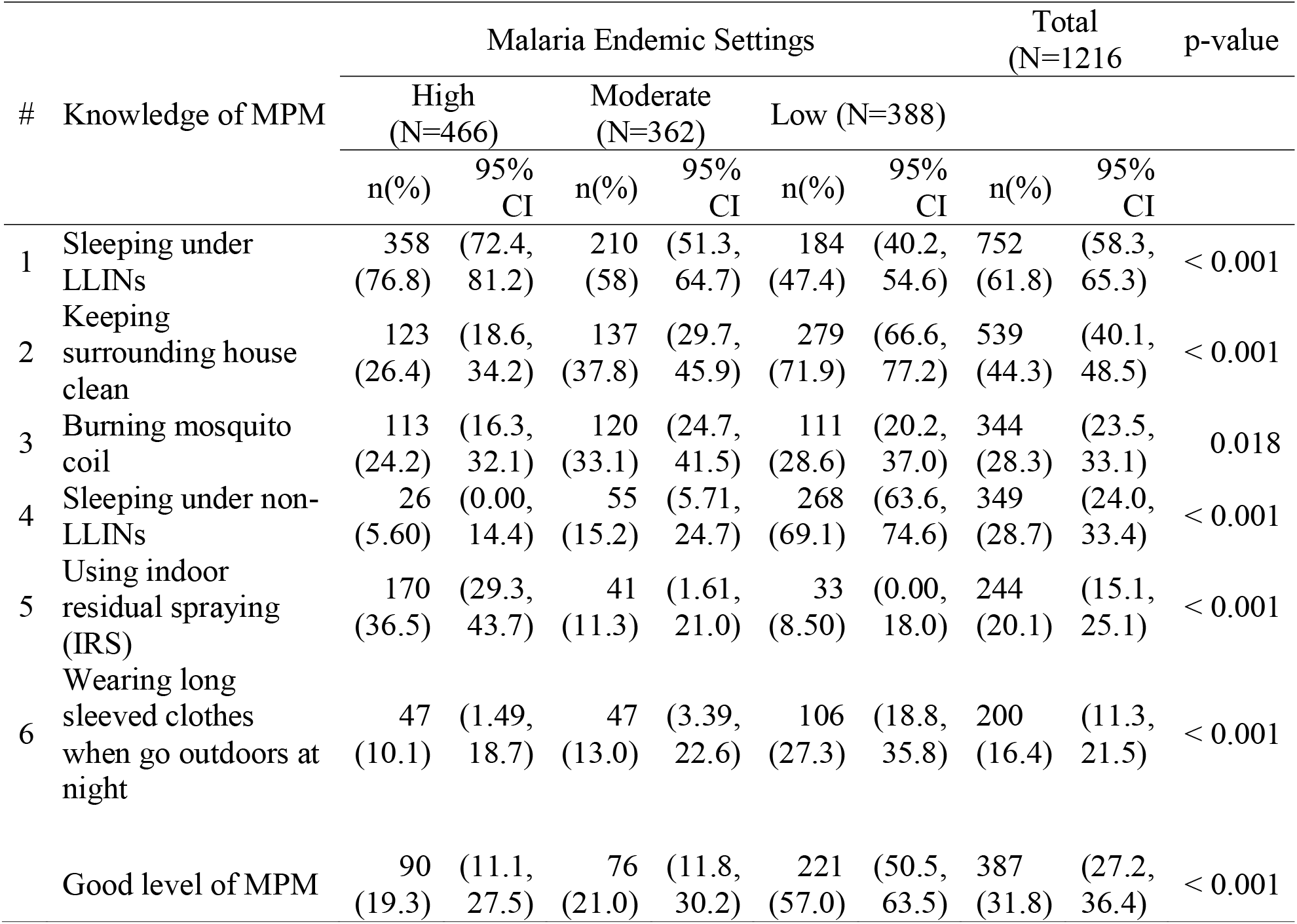
Variation of knowledge of malaria prevention measures of rural adults who have awareness that malaria could be prevented in the East Nusa Tenggara Province, Indonesia (N= 1216)

More than three quarter of participants (76.8%, 95% CI: 72.4 – 81.2) in high MES had knowledge on sleeping under LLINs to prevent malaria. Meanwhile, it was about under half of participants in low MES (45.4%, 95% CI: 40.2 – 54.6). The highest percentage of participants having knowledge on applying non-LLINs to prevent malaria was in low MES at 69.1% with 95% CI: 63.6 – 74.6, whereas it was the lowest in high MES at only 5.6% with 95% CI: 0.00 –14.4. The proportion of participants having knowledge on keeping surrounding house clean to prevent malaria in low MES (71.9%, 95% CI: 66.6 – 77.2) was the highest of other settings. The proportion of participants had knowledge on burning mosquito coil to prevent malaria in low and moderate malaria endemic settings was almost comparable, 28.6% with 95% CI: 20.2 – 37.0 and 33.1% with 95% CI: 24.7 – 41.5 respectively. Meanwhile it was 24.2% with 95% CI: 16.3 – 32.1 in high malaria endemic settings.

### Variation of good level malaria prevention measures knowledge in different malaria endemic settings

The variation of good level of malaria prevention methods knowledge amongst rural adults in different malaria endemic settings was presented in Table 3. The proportion of good level of MPM knowledge between male and female group was not different significantly in all malaria endemic settings. There was a significant different in good level of malaria prevention measures knowledge based on the education level of participants in high (p<0.001) and moderate (p<0.001) malaria endemic settings. In all malaria endemic settings, the good level of malaria prevention measures knowledge was statistically different amongst participants with different socio-economic status and there was a trend that the improvement of good level of malaria prevention knowledge was in line with the increase level of socio-economic status of participants. Regarding the occupation of participants, the highest proportion of good level of malaria prevention measures knowledge was from office workers group with 65.7%, 47.1%, and 41.5% in low, high, and moderate endemic settings respectively.

**Table 3.** Variation of good level malaria prevention measures knowledge among rural adults in different malaria endemic settings by socio-demographic and environmental factors.

| Characteristics | Malaria Endemic Settings |  |  |  |  |  |
| --- | --- | --- | --- | --- | --- | --- |
|  | High |  | Moderate |  | Low |  |
|  | Number<br>at risk | n (%) | Number<br>at risk | n (%) | Number<br>at risk | n (%) |
| **Overall** | **466** | **90(19.3)** | **362** | **76(21.0)** | **388** | **221(57.0)** |
| **Gender** |  |  |  |  |  |  |
| Male | 216 | 48(22.2) | 190 | 45(23.7) | 208 | 111(53.4) |
| Female | 250 | 42(16.8) | 172 | 31(18.0) | 180 | 110(61.1) |
| p-value |  | 0.139 |  | 0.187 |  | 0.124 |
| **Age group** |  |  |  |  |  |  |
| < 30 | 75 | 13(17.3) | 54 | 15(27.8) | 48 | 24(50.0) |
| 30 -39 | 134 | 25(18.7) | 93 | 26(28.0) | 135 | 87(64.4) |
| 40 – 49 | 133 | 29(21.8) | 93 | 14(15.1) | 87 | 56(64.4) |
| 50 – 59 | 66 | 17(25.8) | 79 | 13(16.5) | 76 | 37(48.7) |
| > 60 | 58 | 6(10.3) | 43 | 8(18.6) | 42 | 17(40.5) |
| p-value |  | 0.239 |  | 0.12 |  | 0.013 |
| **Level of education** |  |  |  |  |  |  |
| Primary school or less | 351 | 55(15.7) | 182 | 23(12.6) | 198 | 104(52.5) |
| Secondary school or<br>above | 115 | 35(30.4) | 180 | 53(29.4) | 190 | 117(61.6) |
| p-value |  | <0.001 |  | <0.001 |  | 0.072 |
| **Main occupation** |  |  |  |  |  |  |
| Farmer | 321 | 52(16.2) | 115 | 16(13.9) | 254 | 145(57.1) |
| Housewife | 80 | 19(23.8) | 151 | 25(16.6) | 53 | 23(43.4) |
| Other | 48 | 11(22.9) | 43 | 13(30.2) | 11 | 7(63.6) |
| Office staff | 17 | 8(47.1) | 53 | 22(41.5) | 70 | 46(65.7) |
| p-value |  | 0.008 |  | <0.001 |  | 0.095 |
| **Socio-Economic status** |  |  |  |  |  |  |
| Low | 139 | 11(7.9) | 41 | 3(7.30) | 156 | 76(48.7) |
| Average | 271 | 55(20.3) | 258 | 45(17.4) | 181 | 114(63.0) |
| High | 56 | 24(42.9) | 63 | 28(44.4) | 51 | 31(60.8) |
| p-value |  | <0.001 |  | <0.001 |  | 0.026 |
| **Family size** |  |  |  |  |  |  |
| < = 4 | 190 | 41(21.6) | 260 | 55(21.2) | 183 | 92(50.3) |
| > 4 | 276 | 49(17.8) | 102 | 21(20.6) | 205 | 129(62.9) |
| p-value |  | 0.304 |  | 0.905 |  | 0.012 |
| **The nearest health service** |  |  |  |  |  |  |
| Village maternity posts | 131 | 45(34.4) | 166 | 26(15.7) | 13 | 6(46.2) |
| Village health Post | 34 | 2(5.90) | 56 | 12(21.4) | 127 | 85(66.9) |
| Subsidiary Public Health | 168 | 7(4.20) | 35 | 18(51.4) | 87 | 45(51.7) |
| centres |  |  |  |  |  |  |
| Public Health centres | 133 | 36(27.1) | 105 | 20(19.0) | 161 | 85(52.8) |
| p-value |  | <0.001 |  | <0.001 |  | 0.049 |
| **Distance to the nearest health service** |  |  |  |  |  |  |
| < 1 Km | 165 | 30(18.2) | 177 | 31(17.5) | 118 | 80(67.8) |
| 1 - 2 Km | 84 | 16(19.0) | 121 | 21(17.4) | 123 | 63(51.2) |
| > 2 Km | 217 | 44(20.3) | 64 | 24(37.5) | 147 | 78(53.1) |
| p-value |  | 0.874 |  | 0.002 |  | 0.016 |
| **†HH Income in relation to PMW** |  |  |  |  |  |  |
| < PMW | 431 | 72(16.7) | 327 | 65(19.9) | 324 | 178(54.9) |
| >= PMW | 35 | 18(51.4) | 35 | 11(31.4) | 64 | 43(67.2) |
| p-value |  | <0.001 |  | 0.111 |  | 0.071 |
| **Location of household** |  |  |  |  |  |  |
| Coastal Area | 85 | 20(23.5) | 78 | 18(23.1) | 23 | 4(17.4) |
| Others | 123 | 24(19.5) | 43 | 7(16.3) | 85 | 35(41.2) |
| Hills | 258 | 46(17.8) | 241 | 51(21.2) | 280 | 182(65.0) |
| p-value |  | 0.512 |  | 0.676 |  | < 0.001 |

### Factors associated with good level of knowledge of malaria prevention measures

Factors associated with good level of malaria prevention measures knowledge were presented in Table 4. After controlling all potential confounding variables in multivariate analysis, it was found that in high malaria endemic settings, secondary school or above education level (Adjusted odds ratio (AOR) = 2.37, 95% CI: 1.29 – 4.36); living with high of socio-economic status (SES) (AOR= 3.31, 95% CI: 1.34 – 8.15); living closed to subsidiary public health centre (AOR = 0.09, 95% CI: 0.04 – 0.22), living closed to village health post (AOR = 0.14, 95% CI: 0.03 – 0.64) were significantly associated with good level of malaria prevention measure knowledge. Accordingly, the odds of good malaria prevention knowledge for rural adults in high malaria endemic settings were more two times higher among rural adults with secondary or above education level as compared to those with primary or no education level (AOR) = 2.37, 95% CI: 1.29 – 4.36). Rural adults living with high of socio-economic status had 3 times higher more likely to have good malaria prevention knowledge than those living in low SES (AOR= 3.31, 95% CI: 1.34 – 8.15).

**Table 4.** Factors associated with the good level malaria prevention measures knowledge among rural adults in different malaria endemic settings.

| Characteristics | Malaria Endemic Settings |  |  |
| --- | --- | --- | --- |
|  | High<br>AOR | Moderate<br>AOR | Low<br>AOR |
| **Gender** |  |  |  |
| Male | 1.14 (0.68, 1.91) | 1.46 (0.78, 2.73) | 0.76 (0.48, 1.21) |
| Female | 1.00 | 1.00 | 1.00 |
| **Age group** |  |  |  |
| < 30 | 1.23 (0.39, 3.91) | 1.29 (0.37, 4.46) | 1.82 (0.70, 4.70) |
| 30 -39 | 2.43 (0.88, 6.76) | 1.03 (0.34, 3.08) | 2.85 (1.28, 6.32) |
| 40 - 49 | 2.42 (0.88, 6.65) | 0.70 (0.23, 2.10) | 2.61 (1.14, 5.98) |
| 50 - 59 | 2.55 (0.86, 7.63) | 1.03 (0.34, 3.10) | 1.37 (0.59, 3.15) |
| > 60 | 1.00 | 1.00 | 1.00 |
| **Level of education** |  |  |  |
| Primary school or less | 1.00 | 1.00 | 1.00 |
| Secondary School or above | 2.37 (1.29, 4.36) | 2.66 (1.32, 5.39) | 1.1 (0.67, 1.81) |
| **Main Occupation** |  |  |  |
| Farmer | 1.00 | 1.00 |  |
| Housewife | 1.65 (0.78, 3.50) | 1.35 (0.38, 4.86) |  |
| Other | 0.92 (0.38, 2.19) | 1.76 (0.66, 4.71) |  |
| Office staf | 2.28 (0.71, 7.31) | 2.42 (0.94, 6.26) |  |
| **Socio-Economic Status** |  |  |  |
| Low | 1.00 | 1.00 | 1.00 |
| Average | 1.92 (0.92, 4.00) | 4.46 (1.13, 17.7) | 1.87 (1.14, 3.06) |
| High | 3.31 (1.34,<br>8.15) | 20.5 (4.64,<br>90.8) | 2.52 (1.20,<br>5.30) |
| **Family size** |  |  |  |
| < = 4 |  |  | 0.51 (0.32,<br>0.81) |
| > 4 |  |  | 1.00 |
| **The nearest Health Service** |  |  |  |
| Village maternity posts | 1.00 | 1.00 |  |
| Village health Post | 0.14 (0.03,<br>0.64) | 1.43 (0.59,<br>3.47) |  |
| Subsidiary Public Health centres | 0.09 (0.04,<br>0.22) | 8.35 (3.14,<br>22.2) |  |
| Public Health centres | 0.64 (0.37,<br>1.12) | 0.90 (0.44,<br>1.86) |  |
| **Distance to the nearest health service** |  |  |  |
| < 1 Km | 1.00 | 1.00 | 1.00 |
| 1 - 2 Km |  | 0.94 (0.46,<br>1.94) | 0.42 (0.23,<br>0.77) |
| >=3 Km |  | 3.85 (1.71,<br>8.63) | 0.49 (0.27,<br>0.86) |
| **‡HH Income in relation to PMW** |  |  |  |
| < PMW | 1.00 |  |  |
| >= PMW | 2.12 (0.91,<br>4.95) |  |  |
| **Location of household** |  |  |  |
| Coastal Area |  |  | 0.07 (0.02,<br>0.22) |
| Others |  |  | 0.32 (0.19,<br>0.56) |
| Hills |  |  | 1.00 |

Furthermore, in moderate malaria endemic settings, variables that considerably associated with good level of malaria prevention measure knowledge were secondary school or above education level (AOR = 2.66, 95% CI: 1.32 – 5.39); living with high of socio-economic status (SES) (AOR= 20.5, 95% CI: 4.64 – 90.8); living closed to subsidiary public health centre (AOR = 8.35, 95% CI: 3.14 – 22.2), living more than 2 km from the nearest health facilities (AOR = 3.85, 95% CI: 1.71 – 8.63). Accordingly, the odds of good malaria prevention knowledge for rural adults in moderate malaria endemic settings were more two times higher among rural adults with secondary or above education level as compared to those with primary or no education level (AOR = 2.66, 95% CI: 1.32 – 5.39); Rural adults living with high of socio-economic status had 20 times higher more likely to have good malaria prevention knowledge than those living in low SES (AOR= 20.5, 95% CI: 4.64 – 90.8). The odds of good level malaria prevention knowledge for participants living closed to subsidiary public health centre (AOR = 8.35, 95% CI: 3.14 – 22.2) were eight times higher than those living closed to village maternity post.

Whilst, in low malaria endemic settings, factors such as high SES, having family size less than four, living in coastal area and living closed to the nearest health service were statistically associated with the good level of malaria prevention measures knowledge. Participants living with high of socio-economic status (SES) (AOR= 2.52, 95% CI: 1.20 – 5.30) were nearly three times higher to have good level of malaria prevention measure knowledge than those in low SES. Rural adults living between one and two kilometres from the nearest health service (AOR = 0.42, 95% CI: 0.23 – 0.77) and living more than 2 km from the nearest health service (AOR = 0.49, 95% CI: 0.27 – 0.86) were less likely to have a good level of malaria prevention knowledge than those living less than one kilometres from the nearest health service. Participants living in coastal area (AOR = 0.07, 95% CI: 0.02 – 0.22) were less likely to have a good level of malaria prevention knowledge than those living in hills area.

## DISCUSSION

To our knowledge, this is the first study to investigate the good level of knowledge of malaria prevention measures (MPM) and their associated factors amongst rural adults in different malaria endemic settings of ENTP Indonesia. This study shows that there was statistically different in good level of knowledge of MPM amongst rural adults in high, moderate dan low malaria endemic settings with the lowest was in high malaria endemic settings. This implies that the knowledge related to malaria prevention measures should be scaled up to progress to malaria elimination by 2030. The main factors significantly associated with the good level of MPM knowledge for rural adults in high and moderate settings were the high level of education, and high socioeconomic status. Whilst, in low malaria endemic settings it was high socioeconomic status, distance to the nearest health centres and location of household.

The present study shows that the prevalence of good knowledge of malaria preventative measures in low, moderate and high malaria endemic settings was very low, with 57%, 21% and 19.3% respectively. These findings were lower than reported in rural settings of other nations such as Cameroon (11) and Northwest Ethiopia (12). The low level of malaria prevention methods knowledge of participants in this study might be contributed by the lack of health promotion actions especially related to malaria preventions with the fact that the number of health promotion workers at the level of public health centres in this region was low and their distribution was uneven amongst health centres (34). In addition, nurses and midwifes, beside their main tasks to provide service at health centres, they have to work extra hours without well compensation to educate local people on the significance of MPM particular related to how to apply LLINs appropriately for preventing malaria (35).

This study demonstrated that the proportion of rural adults having knowledge in various methods to prevent malaria was very poor and the discrepancy amongst malaria endemic settings was significant. The proportion of rural adults having knowledge in LLINs which is the main method to prevent malaria adopted by Indonesia government (26) was only common in high malaria endemic settings. This could be understand since the distribution of LLINs in the country was prioritized in high endemic settings (5). Whilst in low malaria endemic settings, the percentage of participants with knowledge in non-LLINs was high. Meanwhile, knowledge on keeping house clean to prevent malaria was only common in low endemic settings. Improving housing condition reduced the density of mosquito in house (36). Whilst other methods of MPM including wearing long sleeved clothes when go outdoors at night, IRS, burning mosquito coil were less knowledgeable by rural adults of ENTP. To boost for malaria elimination efforts, integrated various strategies of MPM tailored with local condition was more advantage than single method (23–25). Having a good understanding of MPM knowledge leads to a greater willingness to practice of MPM (9).

This study further indicates that in all malaria endemic settings, the good knowledge of malaria prevention measures was significantly associated with SES level of participant. The higher the level of SES participants, the higher the level of good knowledge of malaria prevention measures of participants. This finding was in line with study in Southern Ethiopia (12) and Equatorial Guinea (37) indicating that there was a positive correlation between good level of malaria prevention knowledge and SES groups. This might be due to the fact that people from low SES had limited access to multiple source of information (38) including poor access on the internet for health information (39) and they had lower sureness in gaining health information (40).

This study further shows that good level of malaria prevention measures knowledge was significantly associated with education level of participants. This finding was consistent with similar study in other countries such as Malawi (41), Equatorial Guinea (37), and Cameroon (11), indicating that the increasing of malaria prevention knowledge was in line with the improvement of education level of participants. This study discovered that in moderate malaria endemic settings, malaria prevention knowledge of rural adults with secondary or above education level was almost three times higher than those have primary or no education level. Whilst, in high malaria endemic settings, rural adults with secondary school or above education level had more than two times higher more likely to have good malaria prevention knowledge than those had primary or no education level. The reason for this might be that educated people tend to be exposure with multiple sources of information, permitting them to advance their knowledge on MPM (42). These results implies that it is imperative to choose different health communication strategies for educating targeted population about malaria prevention methods tailored to their education background.

The results of this study corroborate with studies in other rural settings to emphasise the power of SES and education in supporting good knowledge of MPM (12,41). As a consequence of good understanding of malaria prevention measures, the behaviour of rural adults might change, leading to increasing good practice of malaria prevention methods in their life. This research provides indication for the low level of good knowledge of MPM in three different malaria endemic settings of rural of ENTP. Improving of knowledge of MPM is critical to reduce the burden of malaria and to progress to malaria elimination. WHO suggests each nation should persist effort to prevent malaria while taking approach to prevent huge impact of COVID-19 [60]. The interruption in delivering malaria prevention tools including insecticide treated nets leads to surge the burden of malaria worldwide (43) and in Indonesia there was an increasing trend of the total number of malaria patients during the COVID-19 pandemic (2). Therefore, well-designed and sustainable approach to expand knowledge of MPM of rural communities would enhance malaria elimination development in this province.

This study has been advantaged by high participation rate of participants in different malaria endemic settings permitting writers to capture the estimation of prevalence of knowledge malaria prevention methods accurately in different settings of rural ENTP. Moreover, data was gathered by visiting household allowing authors to notice the environmental condition including the cleanliness and the usage of mosquito nets in home of participants. However, the authors note some limitations of this study including knowledge of malaria prevention measures based on the self-reported of study participants. It might be diverse if the outcome variables were gained through observation intensively in their daily life. Likewise, the community based cross-sectional study approach was unable authors to infer causal relationship between knowledge of malaria prevention measures and independent variables of the study.

## Conclusions

The good level of malaria prevention measures knowledge in different malaria endemic settings of rural ENTP was very low and the disparity amongst different setting was significant statistically. Higher socio-economic status and education level were considerably associated with the good level of malaria prevention measures knowledge. Therefore, improving MPM knowledge for rural community in moderate and high MES is critical to boost malaria elimination in ENTP. Having high knowledge of MPM would encourage the community to participate in various malaria elimination programs. Targeting the intervention to the low SES and low education level is crucial to boost malaria elimination progress in ENTP Indonesia.

## Data Availability

All data produced in the present study are available upon reasonable request to the authors

